# Unlocking Digital Health: Inequalities in the adoption of a Patient Portal

**DOI:** 10.1101/2025.06.28.25330473

**Authors:** RD Barker, R Gökmen, P Mistry, D Naylor, JT Teo

## Abstract

**Objective:** Digital health apps and patient portals are proposed as part of the drive from ‘analogue to digital’ care for the NHS 10 Year Plan. Without mitigation strategies, digital inequalities could arise as a result and more evidence is needed to understand how to mitigate this.

**Methods:** As part of an equalities impact assessment, a retrospective cross-sectional analysis was conducted examining patient portal activation among patients invited to outpatient appointments at two large south-east London Hospital Trusts between May 1st and November 1st, 2024.

**Results:** 503,688 patients invited to attend outpatients during the study period, 52.7% of patients invited to attend outpatients had activated the patient portal. Availability of email contact details were strongest determinant for likelihood of onboarding (OR 10.86; CI 10.60-11.12). Multivariate logistic regression models showed the following groups were less likely to activate the patient portal. Men (odds ratio 0.84 (CI:0.83–0.85), extremes of age (71-80 years or 11-20 years), those mixed or undefined ethnicity OR 0.58 (CI 0.57– 0.59), black ethnicities OR 0.62 (CI 0.61–0.64) or un-recorded ethnicities OR 0.72 (CI 0.7– 0.74) and those with highest degree of socio-economically deprivation (IMD group 1) OR 0.68 (CI 0.65–0.72).

**Conclusion:** This large scale roll-out of a digital health portal provide empirical evidence of factors which drive digital inequalities for patients of two major London NHS Trusts. The observed disparities across demographic and socioeconomic dimensions and simple reliable digital contact mechanisms highlight the risk that digital healthcare initiatives may inadvertently produce new types of inequalities.

**What is the paper about?:**

1. Were there inequalities in activation of the patient portal, MyChart, in the Apollo programme, by demographic characteristics of the patients? Yes. Table 1
2. Were the observed inequalities attributable to confounding? No. In a multivariate logistic regression, the inequalities persisted across all variables. Figure 1
3. Could the inequalities be explained by differential access to email and mobile phones across the groups? For several variables, adjusting for the presence of email address and mobile phone number attenuated the strength of the relationship with Patient Portal activation. It did not remove the effect for any variable and the relationship with ethnicity was barely affected. Figure 2

## Introduction

Inequality of access is a concern as healthcare systems adopt technological solutions for patient engagement.^1^ Patient portals are digital platforms, such as mobile phone apps and web-sites, which allow individuals to access their medical records, communicate with healthcare providers, and manage appointments. They have become central to healthcare delivery strategies as they can be cheaper to deliver for the healthcare provider and easier to access for the patient. However, these digital tools may inadvertently exacerbate existing health disparities if not implemented with equity considerations.^2^

Understanding patterns of patient portal adoption is particularly relevant given the United Kingdom (UK) National Health Service (NHS) Long Term Plan’s and NHS 10 Year Plan emphasis on digital-first healthcare approaches.^3 4^If certain groups experience limited engagement with patient portals, they may face barriers to accessing information and services, potentially widening existing health inequalities. These concerns align with broader evidence suggesting socioeconomic and demographic factors affect health outcomes and the UK Governments commitment to reducing health inequalities.^5 6^

Previous work studying the UK Health Service’s patient portal, the NHS App, showed geographical differences in uptake and adoption.^7^ The NHS App acts as a patient portal for NHS primary care services but currently has a limited footprint in secondary and tertiary services.

King’s College Hospital NHS Foundation Trust and Guy’s and St. Thomas’ NHS Foundation Trust provide secondary and tertiary health services to a diverse population in London, UK. In 2023, they replaced their Electronic Health Record (EHR) systems and began deployment of a patient portal “tethered” to the new EHR.^8^ This has presented an opportunity to examine patterns of patient portal adoption across a diverse population in the UK.

This research aims to describe demographic and socioeconomic factors associated with adoption of a patient portal with a view to developing targeted interventions that can promote more equitable access to digital health resources across diverse patient populations served by these healthcare institutions.

**Table 1:**
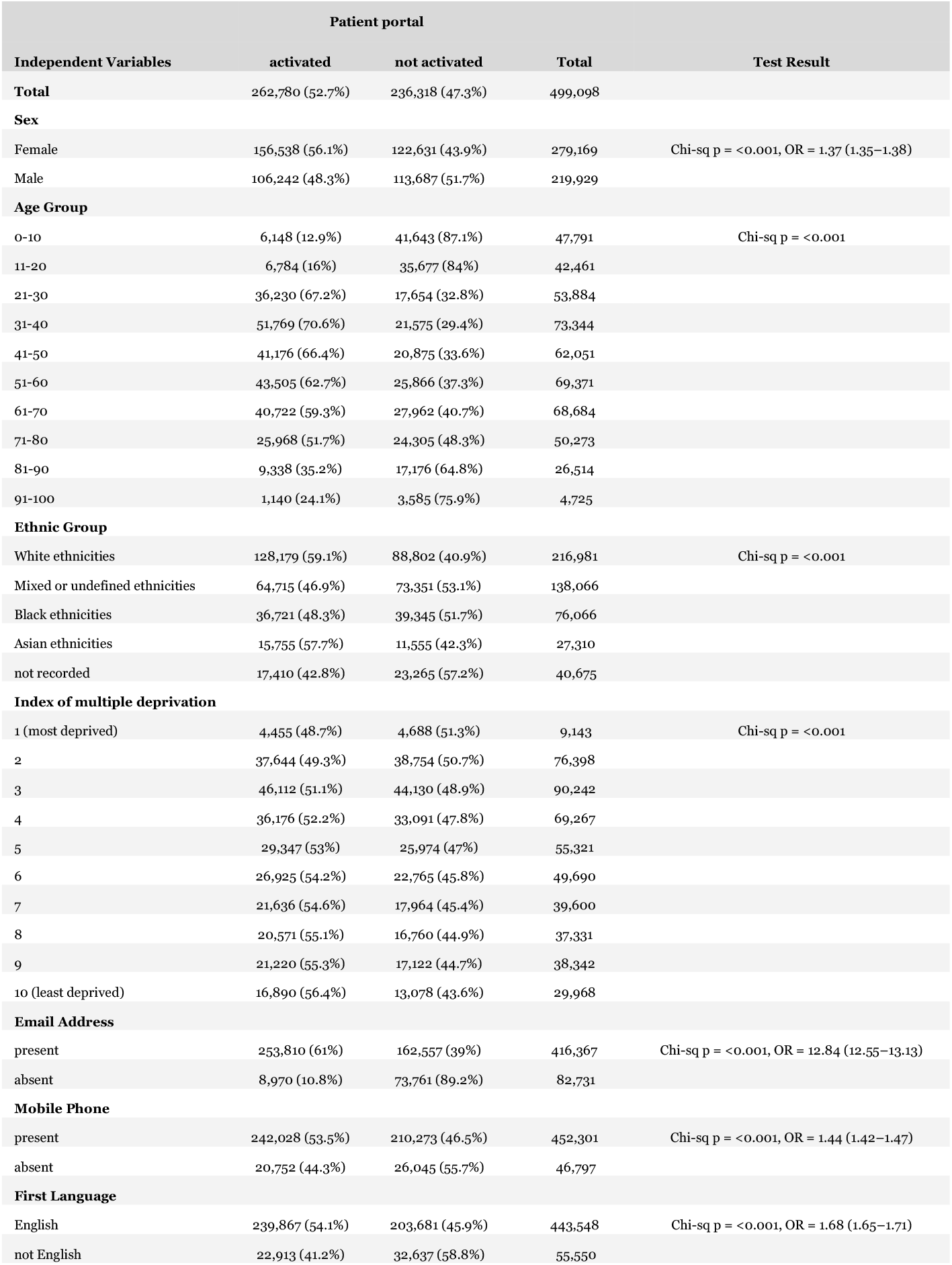
Patient portal activation by Sex, Age group, Ethnic Group, Index of multiple deprivation, availability of email address and mobile phone number and first language among 499,098 patients invited to outpatient clinics across two London teaching hospital systems in south east London, UK, in 2024.

## Methods

### Study Design and Population

A retrospective cross-sectional analysis was conducted examining patient portal activation among patients invited to outpatient appointments at two large south-east London

Hospital Trusts between May 1st and November 1st, 2024. This timeframe was selected to provide a six-month snapshot of portal activation patterns while ensuring adequate sample size for subgroup analyses. At registration patient demographic details were obtained from the NHS spine and by speaking to the patient.^9^ Opportunities to get patients to activate the patient portal were automatically taken at appointment booking, arrival in clinic and at issue of the electronic summary of attendance. Hyperlinks to patient portal sign up were sent by Email and SMS.

Two-step verification of identity was required for a patient to access their records in the patient portal. Due to the cost of SMS messages, the host organisations decided to limit 2- step authentication to email or an authenticator app. A patient without an email address could activate their patient portal account but would have to use an authenticator app to access their personal details.

Before launch, a thorough patient-centred approach was taken to ensure the system reflected diverse experiences and preferences. Feedback from a panel of patients, carers, and community members led to improvements in language, accessibility, and usability. Accessibility standards were independently reviewed and enhancements implemented. After Go-Live focus on inclusion and equity was maintained through annual assessments and continuous community engagement. Helpdesks were established to support users and capture real-time feedback, enabling rapid resolution of issues. Outreach sessions were used to increase user confidence, particularly among older, ethnically diverse, and digitally excluded groups. Volunteers provided in-person support, while a patient panel informed governance and system improvements.

### Data Sources

Data were extracted from the data warehouse, which integrates electronic health record information from the Epic EHR system implemented across both hospital systems. Data was extracted to ensure extraction of one record per patient. The data was correct at the time of data extraction in March 2025. The extraction was performed by the Hospital Trusts’ data analysis team to ensure appropriate data governance, with all analyses conducted on de-identified data.

### Variables and Measurements

The primary outcome measure was patient portal activation status, dichotomised as “activated” or “not activated” (combining status categories of Pending Activation, Inactivated, Non-Standard Status, Patient Declined, and Activation Code Generated but Disabled).

#### Key predictor variables included

- Demographic characteristics:
  ∘ Age (analysed both as continuous and categorized into ten-year age bands)
  ∘ Sex (male/female as recorded in the legal sex field)
  ∘ Ethnicity (collected using NHS standard ethnic categories, analysed both as high-level groupings and detailed categories)^10^
  ∘ First language (English/non-English)
  ∘ Socioeconomic status:
  ▪ Index of Multiple Deprivation (IMD) decile derived from patient postal code linked to Lower Super Output Area (LSOA) data, where 1 represents the most deprived areas and 10 the least deprived
  ∘ Contact information availability:
  ▪ Presence of email address in the electronic health record
  ▪ Presence of mobile phone number in the electronic health record

### Data Analysis

Descriptive statistics were calculated for all variables, stratified by patient portal activation status. All variables were analysed as categorical variables, frequencies and proportions were calculated. Between-group comparisons were conducted using chi- squared tests of independence.

The association between predictor variables and patient portal activation was initially assessed through univariate analyses as described above. Subsequently, multivariate logistic regression models were constructed to evaluate independent associations while adjusting for potential confounding. Two primary multivariate models were developed:

- A base model including demographic variables (age, sex, ethnicity) and socioeconomic status (IMD decile)
- An expanded model additionally incorporating the presence of an email address to evaluate whether contact information availability mediated observed demographic disparities

Adjusted odds ratios (AOR) with 95% confidence intervals were calculated for all predictor variables. Forest plots were generated to visually represent these associations, with reference categories established as female sex, white ethnicity, age group 61-70, and IMD decile 10 (least deprived).

All statistical analyses were performed using R statistical software, with a significance threshold of p<0.05.^11^

### Ethical Considerations

This study forms part of a UK public sector statutory duty for Equalities Impact Assessment (EIA).^10^ It was conducted in accordance with NHS information governance protocols - all data were extracted and analysed in de-identified form, with results presented as aggregated statistics.

## Results

From the 503,688 patients invited to attend outpatients during the study period, complete data was available for 499,098 (99%). Descriptive statistics showed significant differences in patient portal activation across all analysed variables (Table 1).

### Demographic Characteristics and Patient portal Activation

Overall, 52.7% of patients invited to attend outpatients had activated the patient portal at the time of analysis. Women showed higher activation rates than men (56.1% vs. 48.3%, p<0.001). Age groups showed substantial variation in activation, with the highest rates among those aged 31-40 (70.6%) and lowest among children aged 0-10 (12.9%) and elderly patients aged 91-100 (24.1%). White and Asian ethnic groups demonstrated higher activation rates (59.1% and 57.7% respectively) compared to Black ethnic groups (48.3%).

### Socioeconomic Factors and patient portal activation

In univariate analyses, all the variables analysed were related to patient portal activation. A clear socioeconomic gradient was observed in activation rates across IMD deciles.

Patients from the most deprived areas (IMD decile 1) had lower activation rates (48.7%) compared to those from the least deprived areas (IMD decile 10) with activation rates of 56.4% (p<0.001). This gradient was consistent across intermediate deciles, suggesting a robust association between socioeconomic status and patient portal activation.

### Contact Information and patient portal activation

An email address was obtained for 416,367 (83%) of patients and a mobile phone number was obtained for 452,301 (91%). In univariate analysis, the odds ratio for the association between email address availability and patient portal activation was high at odds ratio 12.84 (95% confidence interval 12.55–13.13). Only 3% of those who activated the patient portal did not have an email address. The association with mobile phone was lower, OR 1.44 (CI 1.42–1.47) and English as a first language OR 1.68 (CI 1.65–1.71).

First language also showed an association with activation rates, with English speakers having higher activation rates than non-English speakers (54.1% vs. 41.2%, p<0.001).

### Multivariate analysis of patient portal activation

Multivariate logistic regression models were constructed to identify independent predictors of patient portal activation while adjusting for potential confounding factors.

Figure 1 shows the adjusted odds ratios for sex, age group, ethnicity and deprivation from multivariate logistic regression.

**Figure 1:**
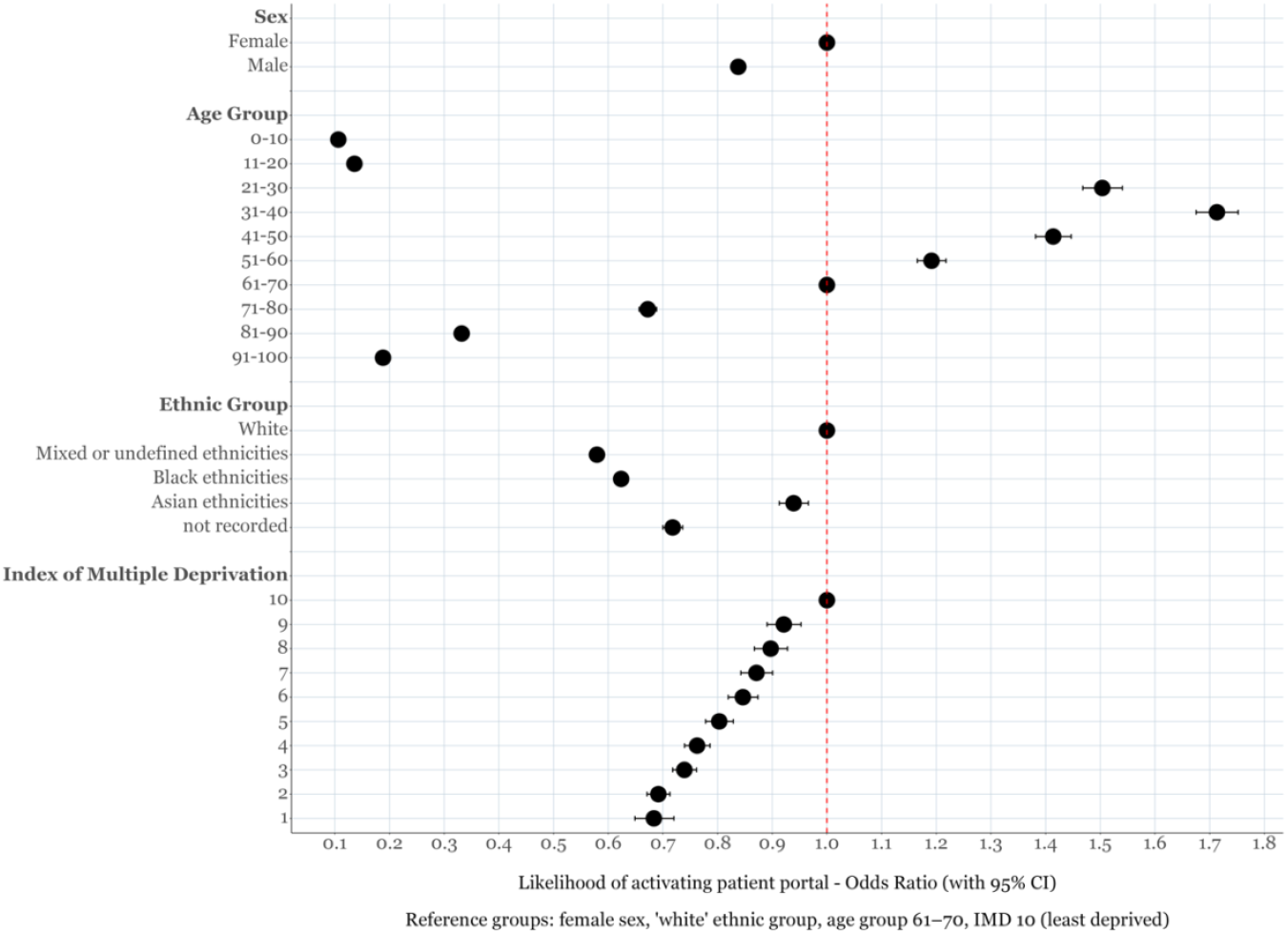
Odds Ratios for adoption of Patient Portal by Sex, Age Group, Ethnic Group and Index of Multiple Deprivation among 499,098 patients invited to outpatients across two London teaching hospital systems between 26 May 2024 and 26 November 2024. Multivariate Logistic Regression.

The odds ratio for the reference groups female sex, ‘white’ ethnic group, age group 61-70, is set at 1. When the odds ratio is less than 1, it indicates that the probability of that group having patient portal activated is less than the reference group. If it is greater than 1, it means that the group is more likely to activate patient portal. Due to large sample sizes, the 95% confidence intervals (the whiskers either side of the dots) are very narrow. When the odds ratios do not overlap, the difference between groups is statistically significant.

It shows that the following groups were less likely to activate the patient portal. Men odds ratio 0.84 (95% CI 0.83–0.85), those of highest age group: 71-80 years OR 0.67 (CI 0.66– 0.69), and lowest ages Age group: 11-20 OR 0.14 (CI 0.13–0.14), those of mixed or undefined ethnicity OR 0.58 (CI 0.57–0.59), black ethnicities OR 0.62 (CI 0.61–0.64) or un- recorded ethnicities OR 0.72 (CI 0.7–0.74) and those more socio-economically deprived e.g. IMD group: 1 OR 0.68 (CI 0.65–0.72).

Figure 2 shows the odds ratios from the same logistic regression model with the addition of having had an email address or mobile phone number recorded in the electronic health record (blue dots).

**Figure 2:**
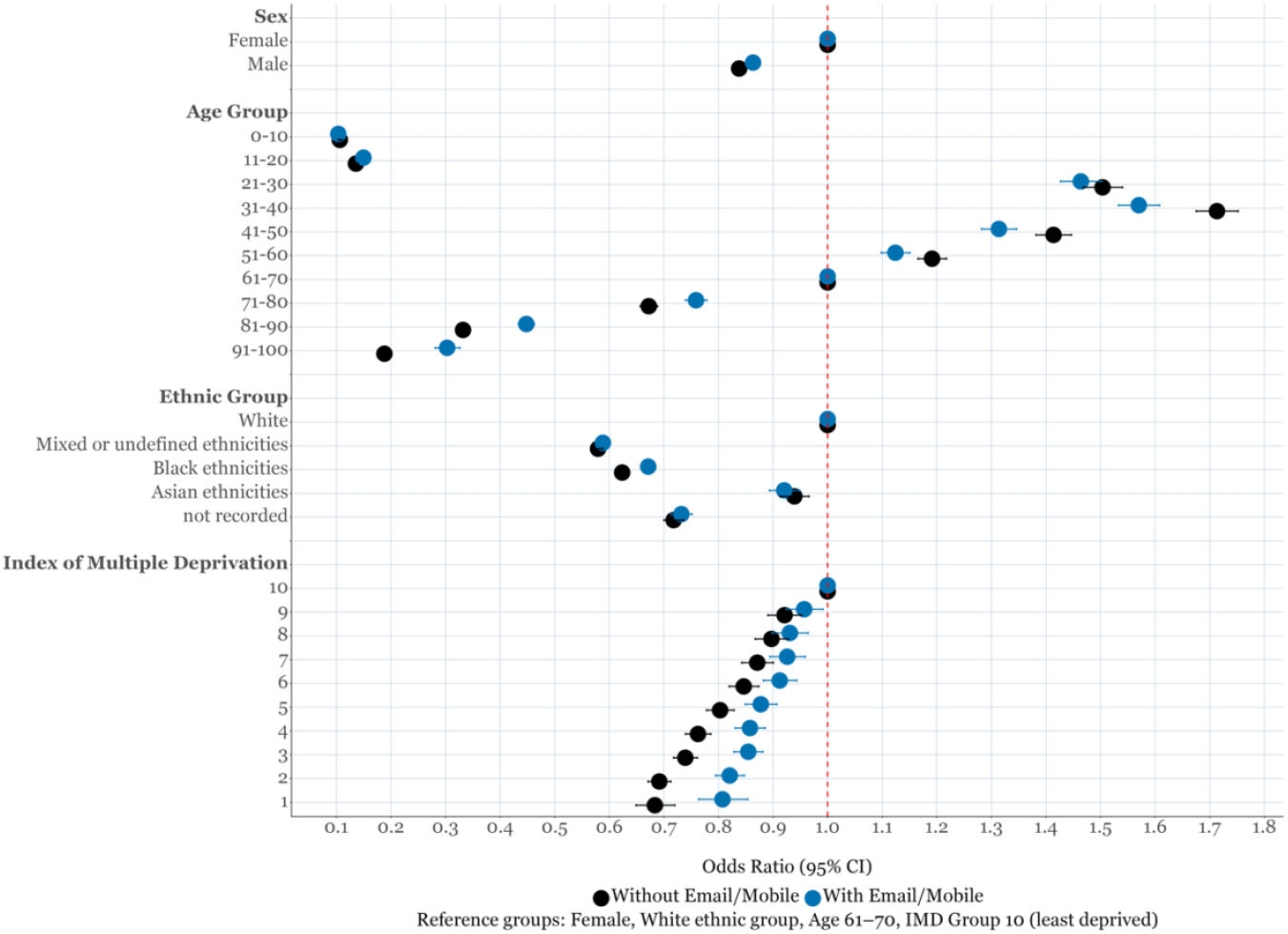
Odds Ratios for **adoption of a patient portal**, by Sex, Age Group, Ethnic Group and Index of Multiple Deprivation, before and after adjusting for the presence of an email address and mobile phone number

The presence of an email address was the strongest predictor of patient portal activation OR 10.86 (CI 10.60, 11.12) with the presence of a mobile phone number less influential OR 1.24 (CI 1.21, 1.27). These are not shown in the figure.

The figure shows that the availability of an email address and mobile phone number had relatively little impact on the effect of sex and ethnicity on patient portal activation. It resulted in a 30 – 40% reduction in the odds ratio associated with deprivation and a 10- 30% reduction in the association with age.

## Discussion

This analysis reveals significant disparities in patient portal activation across demographic and socioeconomic groups. Men, individuals at age extremes (both very young and elderly), black ethnic minorities, and those from areas of higher socioeconomic deprivation demonstrated consistently lower rates of patient portal activation. These patterns persisted in multivariate analyses, suggesting independent effects rather than confounding relationships between these factors. The findings align with previous research on digital health inequalities and raise important concerns about potential widening of healthcare disparities in an increasingly digital NHS environment ^7^.

The strong association between email address availability and patient portal activation represents a key finding with practical implications. As mentioned in the methods, the Trusts decided to limit 2-step authentication to email or an authenticator app. For those without an email address, this may be a step too far and could be an important learning point.

However, it is notable that email address availability did not fully account for the observed demographic disparities shown in figure 2. This suggests that interventions focused solely on collecting email addresses, while necessary, would be insufficient to address the underlying digital inequalities. Interestingly, the distribution of email address availability itself showed similar patterns of inequality across demographic groups, potentially reflecting broader digital literacy and access issues.

The observed ethnic disparities in portal activation warrant particular attention, as they may compound existing inequalities in healthcare access and outcomes. Black ethnic groups consistently showed lower activation rates compared to White and Asian groups, even after adjustment for age, sex, and socioeconomic deprivation. These findings echo research by Zhang et al. on national NHS digital services, which demonstrated similar socioeconomic gradients in adoption of the NHS App across England.^7^ Cultural factors, language barriers, trust in digital health systems, and varying access to digital devices may all contribute to these disparities and require further investigation.

Age-related differences in activation followed a clear pattern, with particularly low rates among paediatric patients, almost certainly attributable to barriers created by parental access issues, and the elderly. The 31-40 years age group showed the highest activation rates (70.6%), while only 24.1% of those aged 91-100 years activated the portal. These substantial disparities suggest that different approaches may be needed for different age groups, with particular attention to digital literacy support for older adults and streamlined proxy access processes for paediatric patients. For working-age adults, barriers may relate more to awareness, competition from other patient portals and perceived utility than to technical capabilities.

Socioeconomic gradients in activation, as measured through IMD deciles, persisted even after adjustment for demographic factors. This suggests that material deprivation itself constitutes a barrier to digital health engagement, potentially through mechanisms including limited internet access, device ownership, digital skills, and competing priorities. These findings raise concerns that as health systems increasingly prioritise digital pathways, socioeconomically disadvantaged patients may face compounded barriers to accessing care and information, potentially widening rather than narrowing health inequalities.^12-14^

The project team implementing the patient portal across our organisations undertook strenuous efforts to proactively prevent the development of inequalities. Informal feedback from patients suggests that the portal is a big leap forward for patients in understanding their care. Despite this, inequalities have clearly emerged.

Healthcare organizations implementing patient portals must develop targeted strategies to address these disparities. Such approaches might include proactive collection of email addresses during registration and clinical encounters; provision of in-person digital support services in clinical settings; culturally tailored education materials; multi- language support; and development of alternative access pathways for digitally excluded populations. Almost all of these have been tried with the patient portal implementation studied in this paper. As the NHS progresses from analogue-to-digital, regular monitoring of portal activation patterns across demographic groups as well as degree of ongoing engagement will be crucial levers to mitigate digital inequalities.

There is an additional unknown risk with the proliferation of digital health tools, websites, apps and portal – multiplicity of digital health apps for different providers and services creating fragmentation of “digital front doors” for patients. It is not realistic for a patient with multimorbidity and multiple healthcare providers to have a different app for each disease, each clinic or each hospital. Digital market forces may create convergence to a few dominant apps, which has its own disadvantages. There is a need for an omnichannel approach to patient portals in a comprehensive healthcare system like the UK NHS. A promising approach is through cross-integration and interoperability as planned with the NHS App to prevent oligopolies of “walled gardens”.^15 16^

Ultimately, digital health tools like patient portal offer significant benefits, but realizing their promise requires deliberate attention to inclusion and equity in their implementation.

## Conclusion

This study provides empirical evidence of significant inequalities in patient portal activation across two major London NHS hospital trusts. The observed disparities across demographic and socioeconomic dimensions—particularly affecting men, black ethnic minorities, older adults, and those from socioeconomically deprived areas—highlight the risk that digital healthcare initiatives may inadvertently widen existing health inequalities. While email address availability emerged as the strongest predictor of patient portal activation, it did not fully explain the observed disparities, suggesting more complex underlying barriers.

UK health care systems have been asked to move from “analogue-to-digital”.^17 18^ These findings underscore the importance of implementing targeted, equity-focused strategies to ensure digital health tools promote rather than hinder equitable healthcare access.^17 18^Healthcare providers should consider developing multi-faceted approaches that address both technical barriers to access and deeper issues of digital literacy, trust, and engagement across diverse patient populations. Regular monitoring of activation patterns across demographic groups should be embedded within institutional equity frameworks to track progress and guide interventions aimed at narrowing the digital divide in healthcare.

## Supporting information

RECORD checklist

## Data Availability

Relevant data in the present work are contained in the manuscript.

## Acknowledgements

Andrew Wilkinson, Aimee Porter Smith and Professor Claire Harrison provided comments and input. JTHT received support from UKRI via the National Institutes of Health Research, Medical Research Council and Health Data Research UK.

## Declarations of Interest

## Contributions

Data collection: DN

Data analysis: RDB, JT

Manuscript drafting: RDB, JT

Critical review: RG, PB

