## Supplemental Table 1 for "Unlocking Digital Health: Inequalities in the adoption of a Patient Portal"

**Supplementary Material**

**Table S1** Breakdown of ethnicity groups (n<100 cells and paired cells are censored)

|  | **My Chart** | |  |
| --- | --- | --- | --- |
| **Ethnicity** | **not activated** | **activated** | **Total** |
| *Unspecified | 12,927 (85.3%) | 2,235 (14.7%) | 15,162 |
| Any Other Ethnic Group | 15,891 (49.6%) | 16,122 (50.4%) | 32,013 |
| Asian or Asian British - Mixed | 269 (42%) | 372 (58%) | 641 |
| Asian or Asian British - Any other Asian background | 3,614 (47.8%) | 3,942 (52.2%) | 7,556 |
| Asian or Asian British - Arab | 586 (51.1%) | 560 (48.9%) | 1,146 |
| Asian or Asian British - Bangladeshi | 1,603 (48.6%) | 1,697 (51.4%) | 3,300 |
| Asian or Asian British - British | 306 (41.3%) | 435 (58.7%) | 741 |
| Asian or Asian British - Caribbean | <100 | <100 | 165 |
| Asian or Asian British - Chinese | 1,410 (34.2%) | 2,710 (65.8%) | 4,120 |
| Asian or Asian British - East African | <100 | 100-200 | 185 |
| Asian or Asian British - Filipino | 216 (24.2%) | 676 (75.8%) | 892 |
| Asian or Asian British - Indian | 3,436 (38.6%) | 5,471 (61.4%) | 8,907 |
| Asian or Asian British - Japanese | <100 | 100-200 | 172 |
| Asian or Asian British - Kashmiri | <100 | <100 | <100 |
| Asian or Asian British - Malaysian | <100 | <100 | 128 |
| Asian or Asian British - Pakistani | 1,492 (43.5%) | 1,935 (56.5%) | 3,427 |
| Asian or Asian British - Punjabi | <100 | 100-200 | 273 |
| Asian or Asian British - Sinhalese | <100 | <100 | <100 |
| Asian or Asian British - Sri Lankan | 201 (39.5%) | 308 (60.5%) | 509 |
| Asian or Asian British - Tamil | <100 | <100 | 130 |
| Asian or Asian British - Unspecified | 543 (50.8%) | 526 (49.2%) | 1,069 |
| Asian or Asian British - Vietnamese | 206 (45.2%) | 250 (54.8%) | 456 |
| Black or Black British - African | 16,868 (51%) | 16,216 (49%) | 33,084 |
| Black or Black British - Any other Black background | 6,767 (56.3%) | 5,249 (43.7%) | 12,016 |
| Black or Black British - British | 796 (42.8%) | 1,062 (57.2%) | 1,858 |
| Black or Black British - Caribbean | 10,990 (51.1%) | 10,515 (48.9%) | 21,505 |
| Black or Black British - Mixed | 275 (50.1%) | 274 (49.9%) | 549 |
| Black or Black British - Nigerian | 1,676 (45.3%) | 2,023 (54.7%) | 3,699 |
| Black or Black British - Somali | 471 (57.5%) | 348 (42.5%) | 819 |
| Black or Black British - Unspecified | 3,044 (52.8%) | 2,718 (47.2%) | 5,762 |
| Central American - Cuban | <100 | <100 | <100 |
| Central American - Mexican | <100 | <100 | <100 |
| Central American - Puerto Rican | <100 | <100 | <100 |
| Mixed - Any other Mixed background | 4,023 (56.9%) | 3,044 (43.1%) | 7,067 |
| Mixed - Asian & Chinese | 100 (33.6%) | 198 (66.4%) | 298 |
| Mixed - Black & Asian | <100 | <100 | 161 |
| Mixed - Black British & White | 251 (47.1%) | 282 (52.9%) | 533 |
| Mixed - Chinese & White | 102 (41%) | 147 (59%) | 249 |
| Mixed - Other/Unspecified | 567 (42.7%) | 761 (57.3%) | 1,328 |
| Mixed - White & Asian | 1,238 (47.6%) | 1,363 (52.4%) | 2,601 |
| Mixed - White & Black African | 1,173 (50%) | 1,171 (50%) | 2,344 |
| Mixed - White & Black Caribbean | 2,118 (47.2%) | 2,373 (52.8%) | 4,491 |
| Not stated/Undefined | 50,081 (54.5%) | 41,813 (45.5%) | 91,894 |
| South American - Bolivian | <100 | 100-200 | 157 |
| South American - Brazilian | 137 (29.5%) | 327 (70.5%) | 464 |
| South American - Columbian | 255 (32.2%) | 538 (67.8%) | 793 |
| White - Albanian | 137 (44.2%) | 173 (55.8%) | 310 |
| White - Any other White background | 12,022 (41.4%) | 16,987 (58.6%) | 29,009 |
| White - Bosnian | <100 | <100 | <100 |
| White - British | 66,028 (41.5%) | 93,111 (58.5%) | 159,139 |
| White - Cornish | <100 | <100 | <100 |
| White - Croatian | <100 | <100 | <100 |
| White - Cypriot (non-specific) | <100 | 100-200 | 240 |
| White - English | 7,019 (38.6%) | 11,180 (61.4%) | 18,199 |
| White - Ex-USSR | <100 | 100-200 | 243 |
| White - Greek | 120 (27.9%) | 310 (72.1%) | 430 |
| White - Greek Cypriot | <100 | 100-200 | 194 |
| White - Gypsy/Romany | <100 | <100 | <100 |
| White - Irish | 1,825 (36.7%) | 3,143 (63.3%) | 4,968 |
| White - Italian | 336 (27.3%) | 893 (72.7%) | 1,229 |
| White - Kosovan | <100 | <100 | <100 |
| White - Mixed | 279 (35.3%) | 512 (64.7%) | 791 |
| White - Northern Irish | <100 | <100 | 111 |
| White - Other European | 1,908 (33.7%) | 3,758 (66.3%) | 5,666 |
| White - Other Ex-Yugoslav | <100 | <100 | <100 |
| White - Polish | 393 (37.2%) | 663 (62.8%) | 1,056 |
| White - Scottish | 216 (35.6%) | 391 (64.4%) | 607 |
| White - Serbian | <100 | <100 | <100 |
| White - Traveller | <100 | <100 | <100 |
| White - Turkish | 300 (40.9%) | 433 (59.1%) | 733 |
| White - Turkish Cypriot | 128 (37.6%) | 212 (62.4%) | 340 |
| White - Unspecified | 957 (41.9%) | 1,327 (58.1%) | 2,284 |
| White - Welsh | <100 | 200-300 | 295 |

**Figure:** Odds Ratios for adoption of Patient Portal by Sex, Index of Multiple Deprivation, Ethnic Group, Age and presence of email address among 499,098 patients invited to outpatients across two London teaching hospital systems between 26 May 2024 and 26 November 2024. Multivariate Logistic Regression.

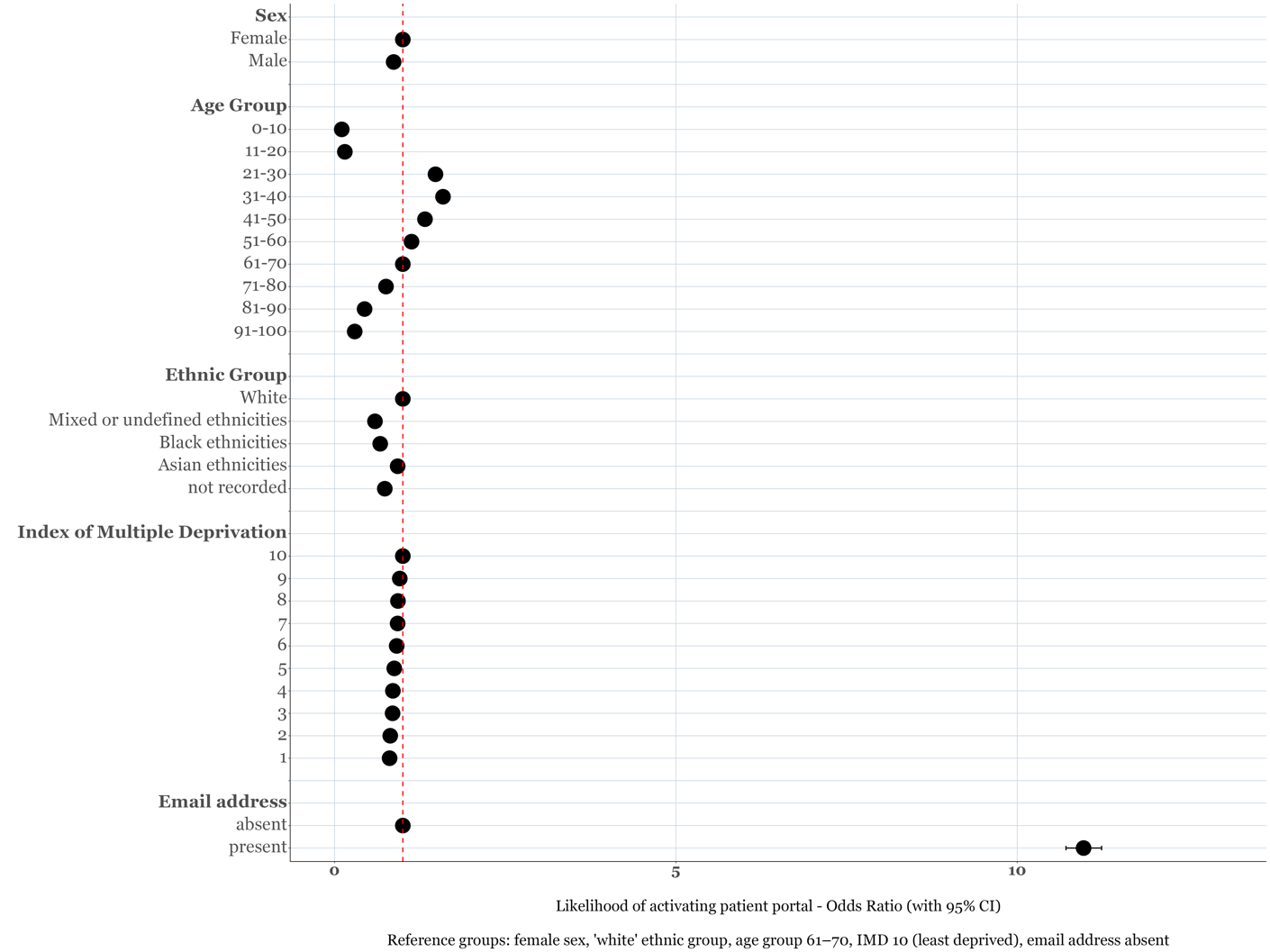

Table 2: the effect of adding Email and Mobile Phone availability on the Odds Ratios for adoption of Patient Portal by Sex, Index of Multiple Deprivation, Ethnic Group, and Age group

| **Variable** | **OR (95% CI) Model without Email/Mobile** | **OR (95% CI) Model with Email/Mobile** | **% Change** |
| --- | --- | --- | --- |
| Sex: Male | 0.84 (0.83–0.85) | 0.86 (0.85–0.87) | -13 |
| Age group: 0-10 | 0.11 (0.1–0.11) | 0.1 (0.1–0.11) | 1 |
| Age group: 11-20 | 0.14 (0.13–0.14) | 0.15 (0.14–0.15) | -1 |
| Age group: 21-30 | 1.5 (1.47–1.54) | 1.46 (1.43–1.5) | -8 |
| Age group: 31-40 | 1.71 (1.68–1.75) | 1.57 (1.53–1.61) | -20 |
| Age group: 41-50 | 1.41 (1.38–1.45) | 1.31 (1.28–1.35) | -24 |
| Age group: 51-60 | 1.19 (1.17–1.22) | 1.12 (1.1–1.15) | -37 |
| Age group: 71-80 | 0.67 (0.66–0.69) | 0.76 (0.74–0.78) | -27 |
| Age group: 81-90 | 0.33 (0.32–0.34) | 0.45 (0.43–0.46) | -18 |
| Age group: 91-100 | 0.19 (0.18–0.2) | 0.3 (0.28–0.33) | -14 |
| Ethnic group: Mixed or undefined ethnicities | 0.58 (0.57–0.59) | 0.59 (0.58–0.6) | -2 |
| Ethnic group: Black ethnicities | 0.62 (0.61–0.64) | 0.67 (0.66–0.69) | -13 |
| Ethnic group: Asian ethnicities | 0.94 (0.91–0.97) | 0.92 (0.89–0.95) | 33 |
| Ethnic group: not recorded | 0.72 (0.7–0.74) | 0.73 (0.71–0.75) | -4 |
| IMD group: 1 | 0.68 (0.65–0.72) | 0.81 (0.76–0.85) | -41 |
| IMD group: 2 | 0.69 (0.67–0.71) | 0.82 (0.79–0.85) | -42 |
| IMD group: 3 | 0.74 (0.72–0.76) | 0.85 (0.83–0.88) | -42 |
| IMD group: 4 | 0.76 (0.74–0.79) | 0.86 (0.83–0.89) | -42 |
| IMD group: 5 | 0.8 (0.78–0.83) | 0.88 (0.85–0.91) | -40 |
| IMD group: 6 | 0.85 (0.82–0.87) | 0.91 (0.88–0.94) | -40 |
| IMD group: 7 | 0.87 (0.84–0.9) | 0.93 (0.89–0.96) | -46 |
| IMD group: 8 | 0.9 (0.87–0.93) | 0.93 (0.9–0.96) | -30 |
| IMD group: 9 | 0.92 (0.89–0.95) | 0.96 (0.92–0.99) | -50 |
| Note: % Change represents the percentage change in the absolute difference from an odds ratio of 1. | | | |

1. Badr J, Motulsky A, Denis J-L. Digital health technologies and inequalities: A scoping review of potential impacts and policy recommendations. *Health Policy* 2024;146:105122. doi: <https://doi.org/10.1016/j.healthpol.2024.105122>

2. Inclusive digital healthcare: a framework for NHS action on digital inclusion. NHS England, . <https://www.england.nhs.uk/long-read/inclusive-digital-healthcare-a-framework-for-nhs-action-on-digital-inclusion>.

3. NHS Digital. Transforming health and care through technology. <https://digital.nhs.uk/about-nhs-digital/our-work/transforming-health-and-care-through-technology>.

4. NHS England. NHS Longterm Plan 2019. 2019. <https://www.longtermplan.nhs.uk/wp-content/uploads/2019/01/nhs-long-term-plan.pdf>.

5. NHS England. Core20PLUS5 (adults) – an approach to reducing healthcare inequalities. <https://www.england.nhs.uk/about/equality/equality-hub/national-healthcare-inequalities-improvement-programme/core20plus5/>.

6. Legislation.gov.uk. Health and Care Act 2022, Section 25, 14Z35 Duties as to reducing inequalities. <https://www.legislation.gov.uk/ukpga/2022/31/section/25/enacted>.

7. Zhang J, Gallifant J, Pierce RL, et al. Quantifying digital health inequality across a national healthcare system. *BMJ Health Care Inform* 2023;30(1) doi: 10.1136/bmjhci-2023-100809 [published Online First: 20231124]

8. British Medical Association. Type 1 opt-out form. <https://bma-mail.org.uk/JVX-7DMBE-36HZKY-4G2VIW-1/c.aspx>.

9. NHS Digital. Spine. <https://digital.nhs.uk/services/spine>.

10. NHS Data Model and Dictionary. Ethnic Category Code 2001. <https://www.datadictionary.nhs.uk/attributes/ethnic_category_code_2001.html?hl=ethnicity>.

11. R Core Team. A Language and Environment for Statistical Computing. <https://www.R-project.org/>.

12. NHS England. Core20PLUS5 – An approach to reducing health inequalities. <https://www.england.nhs.uk/about/equality/equality-hub/core20plus5/>.

13. The King's Fund. What are health inequalities? <https://www.kingsfund.org.uk/publications/what-are-health-inequalities>.

14. Public Health England. Place-based approaches for reducing health inequalities. <https://www.slideshare.net/PublicHealthEngland/placebased-approaches-for-reducing-health-inequalities>.

15. NHS App to be integrated with Epic EPR in 2025. <https://www.digitalhealth.net/2025/03/nhs-app-to-be-integrated-with-epic-epr-in-2025/>.

16. NHS England. How to integrate with the NHS App. <https://digital.nhs.uk/services/nhs-app/how-to-integrate-with-the-nhs-app>.

17. NHS England. 2025/26 priorities and operational planning guidance. <https://www.england.nhs.uk/long-read/2025-26-priorities-and-operational-planning-guidance/>.

18. Department of Health and Social Care. Road to recovery: the government's 2025 mandate to NHS England. <https://www.gov.uk/government/publications/road-to-recovery-the-governments-2025-mandate-to-nhs-england/road-to-recovery-the-governments-2025-mandate-to-nhs-england>.
